# Research on the Emotions of Uninfected People during the COVID-19 Epidemic in China

**DOI:** 10.1101/2022.03.22.22272730

**Authors:** Gang Chen, Juan-Juan Yue

## Abstract

**Background:** The negative emotions induced by the ongoing coronavirus disease 2019 (COVID-19) epidemic are affecting people’s health. In order to identify emotional problems and promote early intervention to reduce the risk of disease, we studied the emotional states of Chinese people during the epidemic.

**Method:** We adopted the Automated Neuropsychological Assessment Metrics mood scale and prepared an online questionnaire. Then, we conducted an exploratory factor analysis of the effective responses of 567 participants from 31 provinces and cities in China. Finally, we analyzed the characteristics of the distribution of different types of emotions and compared them via several statistical methods.

**Results:** The original scale was modified to have six dimensions that yielded reliable internal consistency values ranging from 0.898 to 0.965 and explained 74.96% of the total variance. We found that a total of 33.9% of respondents felt negative emotions more strongly, were less happy and had less energy than other respondents (p<0.001). People with these traits had relatively serious emotional problems and were typically over 60 years old, doctoral degree holders, enterprise personnel and residents in an outbreak area.

**Conclusion:** Thirty-three percent of people without COVID-19 had emotional problems. Psychotherapy should be provided as early as possible for people with emotional problems caused by the epidemic, and the modified scale could be used to survey the public’s mood during public health events to detect problems and facilitate early intervention.

## Introduction

Since December 2019, novel coronavirus disease 2019 (COVID-19) has spread from Wuhan, Hubei Province, China. This sudden public health event has led to uncertainty and should be recognized as a stressful situation that can affect not only people’s physical health but also their psychosocial well-being (Slavich, 2016). According to reports, in 2003, during the severe acute respiratory syndrome (SARS) epidemic, there was a 30% increase in suicide in those aged 65 years and older, approximately 50% of recovered patients remained anxious after recovery, and 29% of healthcare workers showed signs of emotional distress (Nickell et al., 2004; Tsang et al., 2004; Yip et al., 2010). Many researchers predicted that the COVID-19 pandemic would cause emotional problems, such as anxiety, depression, self-harm, and suicide attempts, across age groups (Holmes et al., 2020; Lima et al., 2020; Yang et al., 2020). This was predicted to be the result of an increase in social isolation and loneliness caused by the COVID-19 pandemic (Holmes et al., 2020). Some studies have shown that different populations have suffered moderate or severe psychological impacts during the COVID-19 epidemic (Cao et al., 2020; Lai et al., 2020; Wang et al., 2020).

Mood states reflect the prior expectations about precision that nuances (emotional) fluctuations in confidence or uncertainty (Clark et al., 2018). The brain seeks to maintain its physiological (and psychological) equilibrium despite constantly changing internal and external environments, which renders conditions predictable and minimizes or even resolves uncertainty or surprise (Friston, 2009). When people are constantly in stressful situations, stressors can permanently affect key neurobiological systems and disrupt their structure and/or function; this can lead to the development of mood disorders (Clark et al., 2018), which are associated with poor quality of life and increased mortality. For example, major depressive disorder is the second leading cause of disability globally (Ferrari et al., 2013).

Currently, little is known about the emotional state of the general public. This study aims to assess the mood states of uninfected persons using modified scales, present the prevalence of emotional problems and probable reasons, and identify the characteristics of people who are prone to certain emotional problems or severe emotional problems. These findings may draw the government’s attention to the emotional problems of the public and may assist health care professionals in developing detailed plans for psychological interventions in the continuing COVID-19 epidemic in China and different parts of the world.

Many methods of assessing mood have been developed. The Automated Neuropsychological Assessment Metrics (ANAM) is a computer-based measure of cognitive performance originally developed by the U.S. Department of Defense to monitor changes in performance in healthy individuals experiencing environmental challenges (Reeves et al., 1992). Subsequently, the ANAM has been used extensively to measure cognition for many clinically and non-clinically relevant applications, such as detecting deficits in patients with multiple sclerosis, systemic lupus erythematosus, and Parkinson’s disease, and to measure human performance (Kane et al., 2007; Vincent et al., 2012).

The ANAM system combines mood, sleep and cognitive testing to monitor neuropsychology; it was first introduced in the early 1990s and has undergone a number of revisions (Vincent et al., 2012). The mood scale (AMS) of version 4.0 of the ANAM (ANAM4) contains 7 dimensions, namely, vigour, restlessness, depression, anger, fatigue, anxiety, and happiness, and each domain includes 6 items (Johnson et al., 2008). Compared with other popular mood scales, the AMS has been shown to have excellent test–retest reliability and internal consistency (Johnson et al., 2008). However, the AMS has not been used to assess the mood states of Chinese people during public health events. The scale can be adjusted based on the respondent and situation.

## Materials and methods

### Instruments and measurements

The AMS was translated into Chinese and was modified by two specialists with experience working overseas. Then, we prepared the questionnaire, “The investigation of healthy people’s mood states during coronavirus disease 2019”, with 42 items assessed on a 7-point Likert scale (1=not very strong, 7=very strong). The overall reliability and the reliability of each dimension of the self-rating scale were excellent, with a Cronbach’s alpha coefficient of 0.924 for the total scale and coefficients ranging from 0.898 to 0.965 for the six subscales (Table 1).

**Table 1.** Cronbach’s alpha scores.

|  | Total | Manic depression | Happiness | Restlessness | Vigour | Fatigue | Anxiety |
| --- | --- | --- | --- | --- | --- | --- | --- |
| Cronbach’s $\alpha$ | 0.924 | 0.965 | 0.940 | 0.926 | 0.898 | 0.905 | 0.942 |

### Participants and data collection

An anonymous survey with the prepared questionnaire was administered online from March 9, 2020, to March 16, 2020, and only one survey response could be submitted from a single IP address. A total of 573 persons from 31 provinces and cities in China voluntarily participated in the survey. Six returned questionnaires were excluded, two because they were from COVID-19 patients and four because they were from overseas individuals. Therefore, the effective sample size was 567, with an effective response rate of 99%. The sample size conformed to the principle that the ratio of items to subjects should be 1:5 and that the total number of respondents should be no less than 100 (Gorsuch, 1983). The sample size was determined before the data analysis was performed.

### Ethics statement

The study was conducted in accordance with the Declaration of Helsinki and was approved by the Ethics Committee of the author’s University. All experiments were performed in accordance with relevant guidelines and regulations. Informed consent was obtained from all subjects.

### Data analysis

Exploratory factor analysis (EFA) was used to explore the latent construct explaining the relationships among the measured variables. Principal component analysis (PCA) was used for exploration of the theory of mood states rather than for data reduction purposes. The scores of the items in each subscale were summed to generate scores for the domains of manic depression, happiness, restlessness, vigour, fatigue and anxiety, and the average score of each person for each domain was used in the analysis. The data were analysed using SPSS 13.0 (SPSS, Inc., Chicago, IL). The demographic data were analysed using descriptive statistics, and product-moment correlation analysis was used to examine the relationships among the six domains and the relationship between different emotions and the number of confirmed cases or deaths. Independent sample T-tests and ANOVAs were used to test differences in people’s negative emotions in the four negative emotion domains, and P<0.05 indicated statistical significance.

## Results

### Demographic information of the participants

A total of 567 participants (301 men and 266 women) with a mean age of 38.77 ± 11.24 years from 31 provinces and cities in China were investigated. The participants were mainly from Sichuan (24.0%), Chongqing (22.2%) and Hubei (13.9%) Provinces. The majority of participants had bachelor’s degrees (39.9%), followed by master’s degrees (22.2%) and doctoral degrees (17.3%). The respondents were mainly frontline healthcare workers (24.0%) fighting COVID-19, employees of companies (20.3%) and teachers (15%). The demographic information is shown in Supplementary Table 1.

### EFA results

PCA with varimax rotation was conducted on 42 items. The Kaiser-Mayer-Olkin (KMO) measure found that the sample size was adequate for the analysis (KMO=0.96). Bartlett’s test of sphericity (χ^2^(861)=25112.896, p<0.001) indicated that the correlation coefficients among all of the items were sufficiently large to perform EFA.

EFA provided a simple six-factor construct with 42 items that explained 74.96% of the total variance. The eigenvalues and percentages of the explained common variance are shown in Table 2. All the items loaded on the principal components were greater than 0.5, and no items needed to be discarded (Table 3). Because the eigenvalues were above 1.00 both before and after the rotation, these six common factors could be selected and were used to generate the structure of the uninfected people’s mood states. The six factors were named manic depression, happiness, restlessness, vigour, fatigue and anxiety according to the items they contained (Table 3). These factors were significantly correlated (p<0.001); manic depression, restlessness, fatigue, and anxiety were highly correlated (r>0.70), and restlessness and fatigue were moderately related (0.4≤r=0.596≤0.7) (See Supplementary Table 2).

**Table 2.**
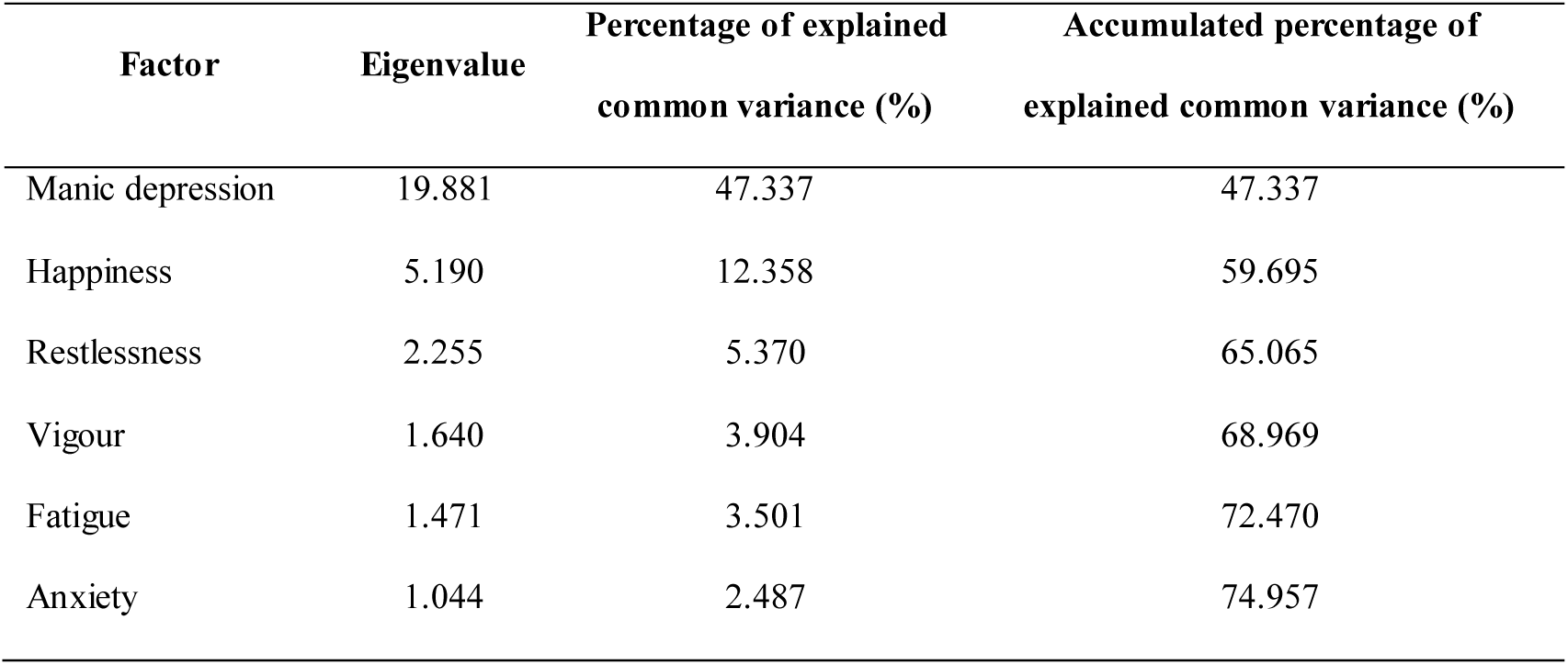
Eigenvalues and percentages of explained common variance.

**Table 3.**
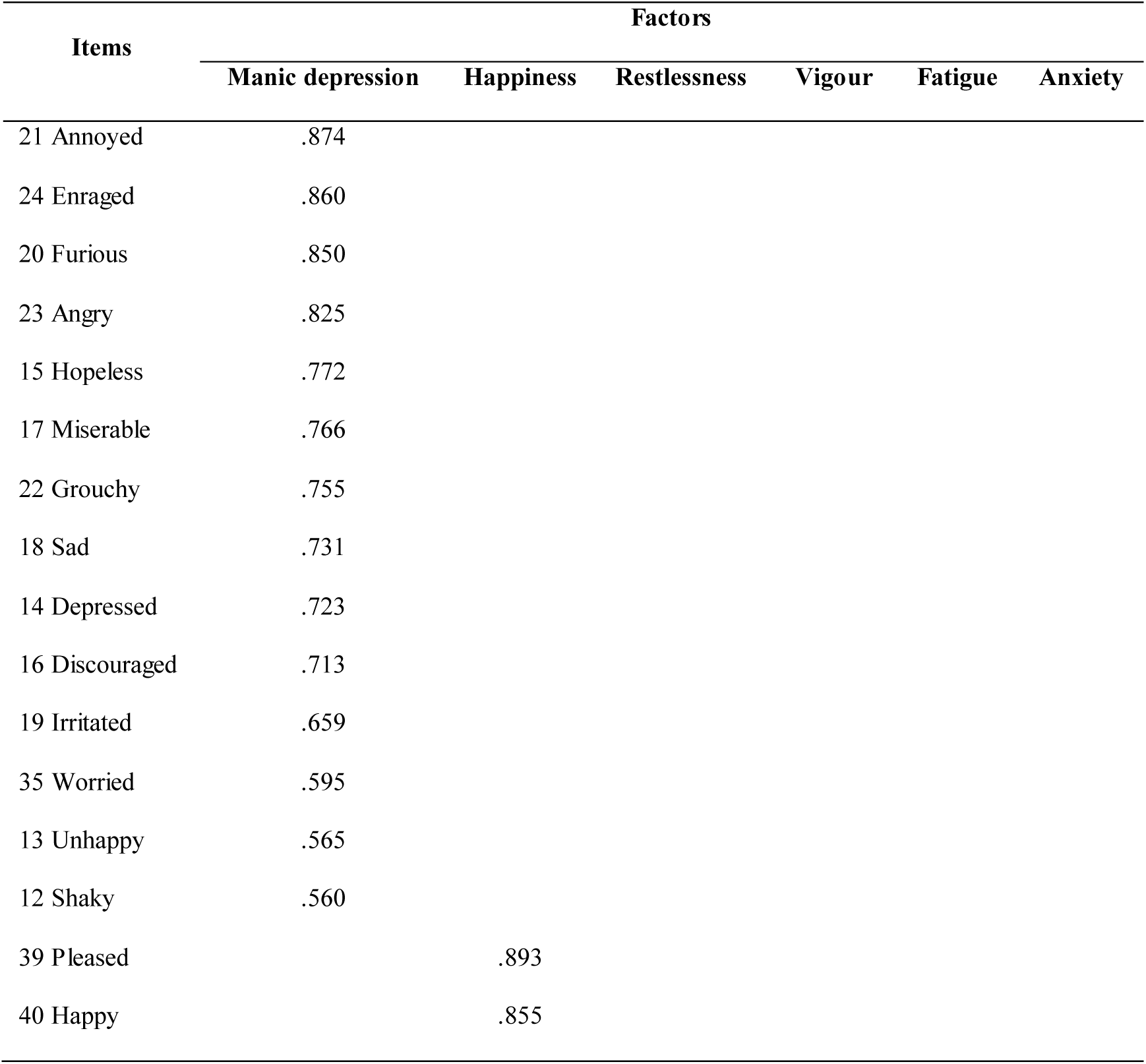
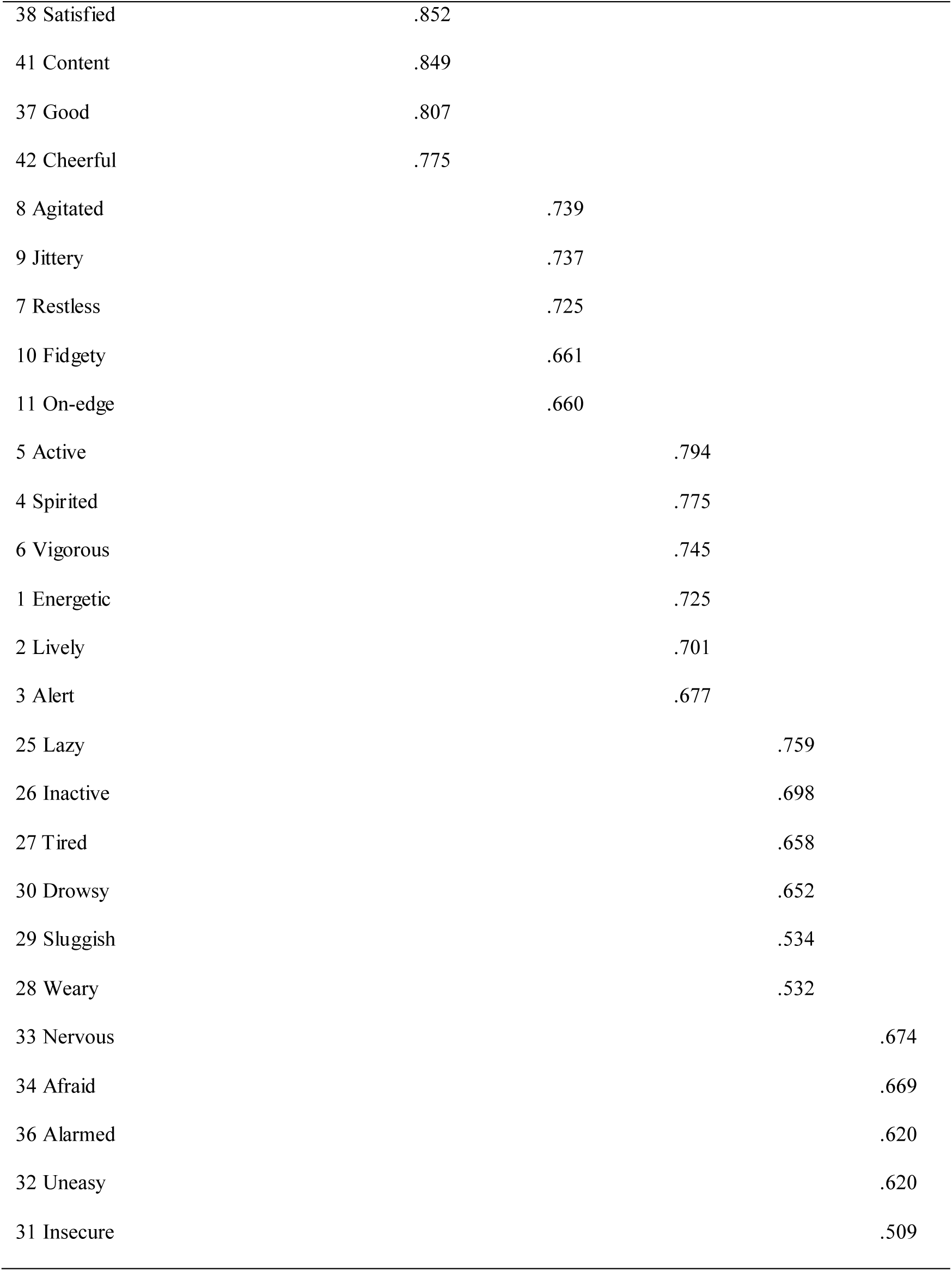
Principal component analysis with varimax rotation and Kaiser normalization.

### Participants’ mood states

Most people were in a good mood. The highest mean score was found for vigour (m=4.91, sd=1.11), followed by happiness (m=4.16, sd=1.45), fatigue (m=2.87, sd=1.39), restlessness (m=2.59, sd=1.41), anxiety (m=2.44, sd=1.43), and manic depression (m=2.11, sd=1.20). However, 33.9% of uninfected people (n=192) reported at least one type of negative emotion. Those who reported at least one type of negative emotion felt negative emotions more strongly, were less happy and had less energy than those who did not report any negative emotion (p<0.001) (Figure 1a). The perception of fatigue was the strongest (4.25±1.19) and that of manic depression was the weakest (3.26±1.26) among the four negative emotions. We divided people who experienced negative emotions into four groups, namely, manic depression, restlessness, fatigue and anxiety, according to whether their average scores in each domain were equal to or greater than 4 points; a score of 4 points or greater indicated that they were unsure whether they felt the emotion or felt the emotion at least to some extent. In total, 24% felt fatigued, 20.3% felt restless, 18.2% felt anxious and 9.9% felt manic depressive. People in these groups reported significantly more negative emotions than others (p<0.001) (Figure 1b).

**Figure 1.**
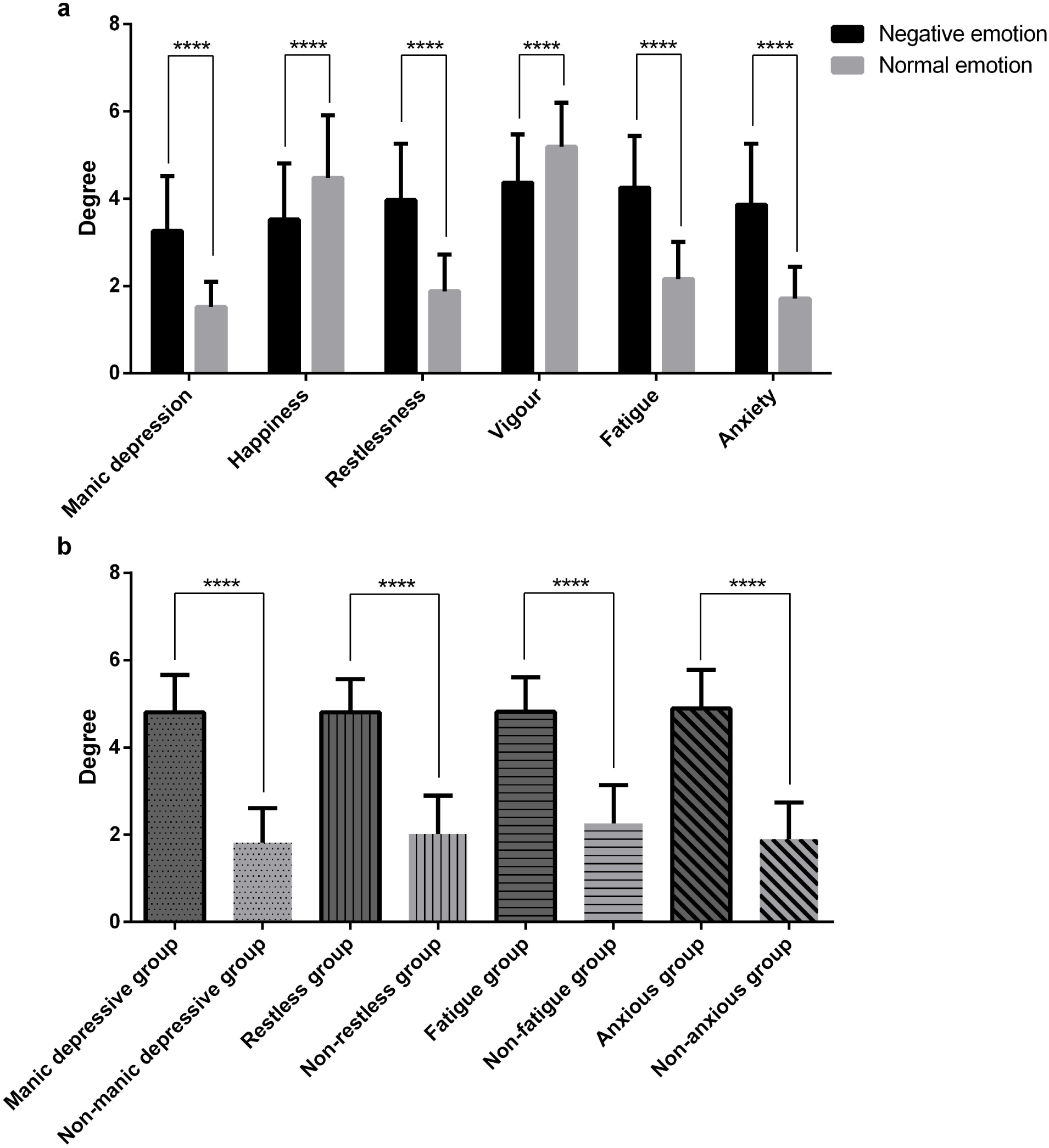
Comparison of the perceived degree of emotions among different groups. a. Difference in the perception of six emotions among the negative emotion group and normal emotion group. b. Comparison of negative mood perception between groups. ****p<0.0001

A slightly higher percentage of women (34.2%) than men (33.6%) experienced negative emotions (See Supplementary Table 3). Women mainly felt fatigued, and a greater percentage of women (26.3%) felt fatigued than men (21.9%) (Figure 2a). The proportions of men and women with other negative emotions were similar. There was no significant difference between the sexes regarding the perception of each emotion.

**Figure 2.**
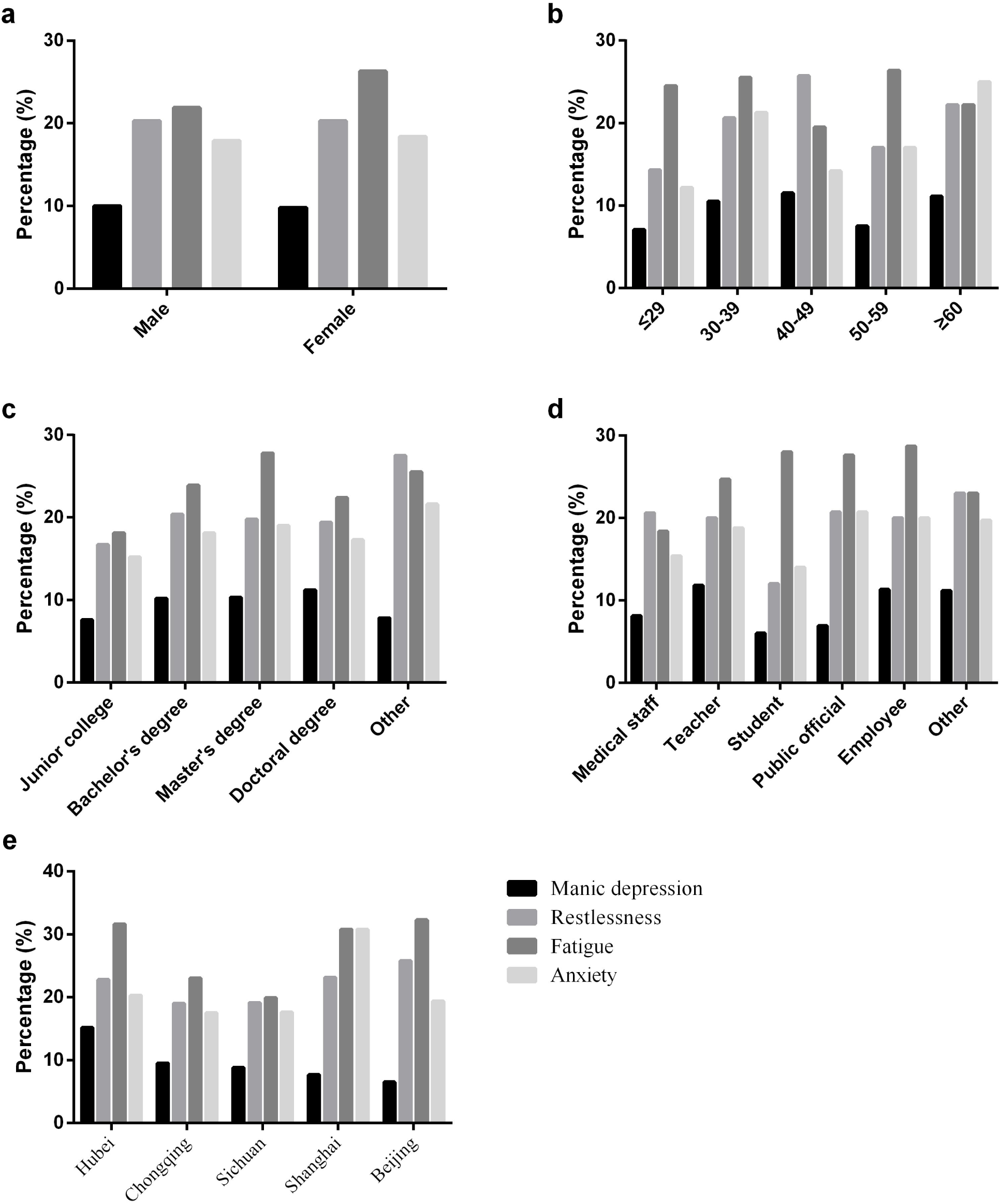
The distributions of the perception of negative emotions among different groups. a. Male and female. b. Different age groups. c. Different educational background groups. d. Different occupation groups. e. Different location groups

The average age of the persons who experienced negative emotions was approximately 39 years old (See Supplementary Table 3). People under the age of 29 were the least likely of all age groups to report negative emotions (31.6%). People over 60 years mainly felt anxious (25%), those between 50 and 59 years old felt fatigued (26.4%), and those aged 40 to 49 years were most likely to feel manic depressive (11.5%) and restless (25.7%) (Figure 2b). People over 60 years old felt manic depressive emotions significantly more strongly than those between 30 and 39 years (p=0.023) and those between 50 and 59 years (p=0.044) (Figure 3a), and they were more restless (p=0.045) (Figure 3b) and fatigued (p=0.007) (Figure 3c) than those under 29 years. Meanwhile, people between 40 and 49 years felt much more fatigued (p=0.018) (Figure 3c) and anxious (p=0.046) (Figure 3d) than those under 29 years. There were no significant differences among the other age groups.

**Figure 3.**
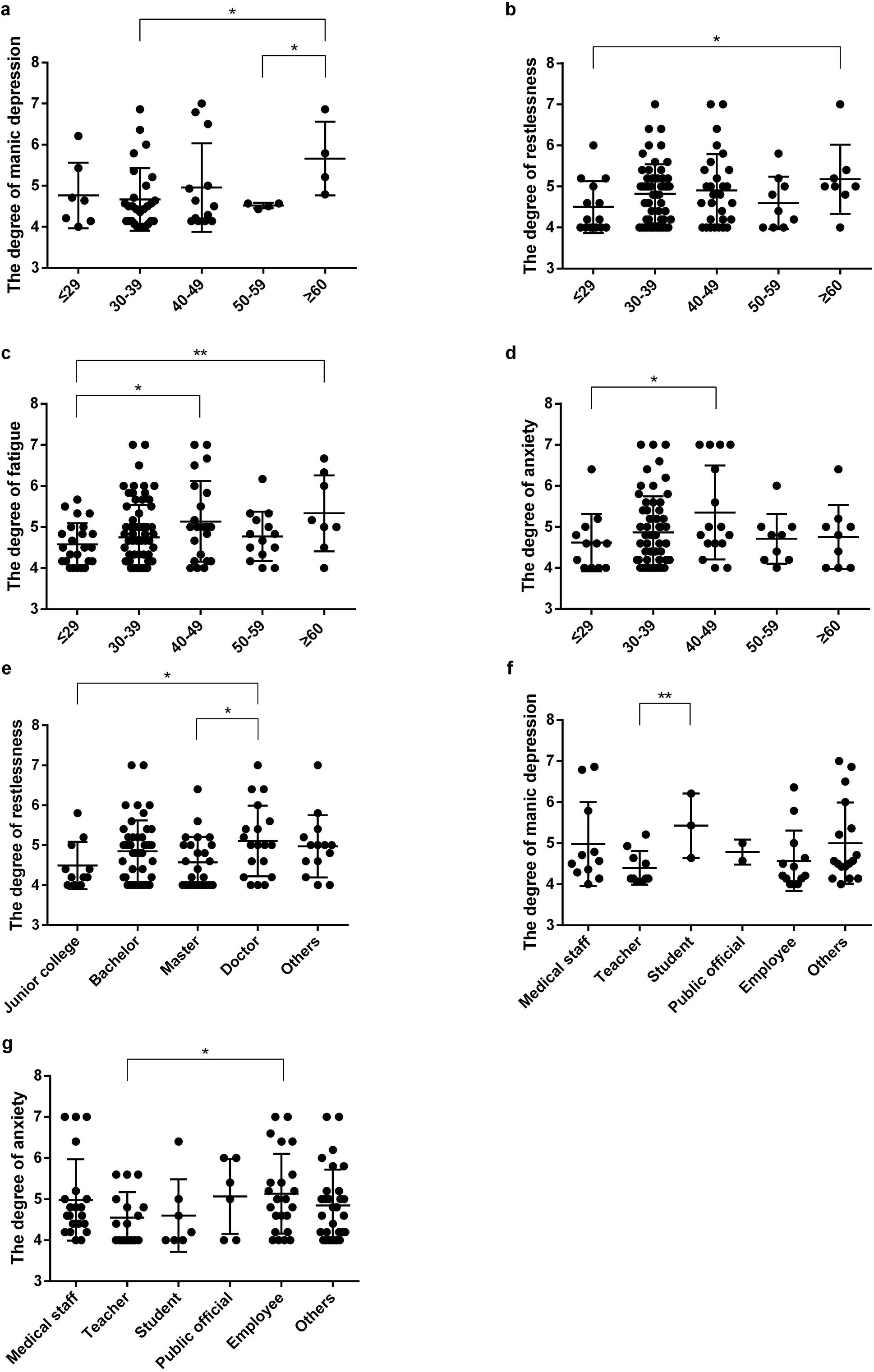
Comparison of the perceived degree of negative emotions among different groups. a. The difference in the perception of manic depression among age groups b. The difference in the perception of restlessness among age groups c. The difference in the perception of fatigue among age groups d. The difference in the perception of anxiety among age groups e. The difference in the perception of depression among occupation groups f. The difference in the perception of anxiety among occupation groups g. The difference in the perception of restlessness among educational background groups * p<0.05,* *p<0.01

People with less than a college degree showed the highest rates of negative emotions (41.2%) (See Supplementary Table 3), and the percentages of those who experienced restlessness (27.5%) and anxiety (21.6%) in that group were higher than those in the other groups (Figure 2c). The proportion of people who experienced fatigue was highest in the group of people with master’s degrees (27.8%) (Figure 2c). There was no significant difference in the perception of negative emotion among people with different educational backgrounds, except that people with doctoral degrees felt more restless than those with master’s degrees (p=0.024) and junior college degrees (p=0.0497) (Figure 3e). Meanwhile, the mean scores for manic depression (5.07±1.19) and anxiety (5.21±1.17) were the highest in the doctoral degrees group and no significant difference in these scores between all groups was found (See Supplementary Table 4).

Among the occupational groups, the group of healthcare workers had the lowest proportion of people who reported experiencing negative emotions (30.1%) (See Supplementary Table 3). The student group had the lowest proportions of participants reporting manic depressive symptoms (6.0%), restlessness (12.0%) and anxiety (14.0%), and 28% of students reported fatigue (Figure 2d). However, there were no significant differences in the degree to which different occupational groups felt the same emotions, except that students felt manic depressive significantly more than teachers (p=0.009) (Figure 3f) and employees were more anxious than teachers (p=0.042) (Figure 3g).

In the survey, the five provinces and cities with the highest proportions of participants reporting negative emotions were Beijing (45.2%), Shanghai (42.3%), Hubei (36.7%), Chongqing (31.7%) and Sichuan (31.6%) (See Supplementary Table 3). Hubei Province had the highest proportion of people who reported feeling manic depression (15.2%) (Figure 2e). People from Hubei Province had the highest mean scores for the perception of manic depression (4.87±0.90) and anxiety (5.01±1.10); however, there were no significant differences in the degree of the perception of all negative emotions among people from different regions (See Supplementary Table 4). In addition, we found that the perceived degrees of manic depression (p<0.01), fatigue (p<0.01) and anxiety (p<0.05) were positively correlated with the number of confirmed cases or deaths. However, the perceived degree of vigour was negatively correlated with the number of confirmed cases or deaths (p<0.01) (See Supplementary Table 5).

## Discussion

Because the government effectively controlled the epidemic, most people perceived positive emotions, with the strongest perceptions of vigour (m=4.91, sd=1.11) and happiness (m=4.16, sd=1.45). However, 33.9% of people still showed significant signs of manic depression, restlessness, fatigue or anxiety (p<0.001). These ratios are lower than those reported in a previous study, which reported that 53.8% of respondents showed psychological impacts within the first two weeks of the COVID-19 outbreak in China (Wang et al., 2020). These people mainly experienced fatigue, followed by restlessness, anxiety and manic depression, significantly more than others who did not show psychological impacts (p<0.001). The four kinds of negative emotions were significantly correlated with each other. In other words, these emotions are often experienced at the same time or set off a chain reaction.

Mental fatigue refers to the feeling that people may experience after or during prolonged periods of cognitive activity (Boksem & Tops, 2008). Mental fatigue can cause decreased cognitive task performance and impaired endurance and make it difficult to maintain an adequate level of performance on tasks (van Cutsem et al., 2017). Mental fatigue might also increase subjective feelings of tiredness and decrease vitality and motivation (Boksem & Tops, 2008). The results of this study were consistent with this expectation, and fatigue was moderately associated with reduced vigour (r=-0.504, p<0.01). Fatigue was common among the surveyed people and corresponded with a desire to abandon certain behaviours that were not perceived as changing the situation during the prolonged epidemic, consistent with the theory of effort/reward imbalance (Boksem & Tops, 2008). Restlessness refers to a state of poorly organized and aimless motor activity stemming from physical or mental unease and can be described as “finding or affording no rest; uneasy, agitated, constantly in motion, fidgeting, etc (Sachdev & Kruk, 2016).” Restlessness is usually observed in individuals with depression and anxiety (Sachdev & Kruk, 2016); this was supported by the results of this study, which showed strong positive correlations between restlessness and both manic depression and anxiety (r=0.742, r=0.724, p<0.01). Restlessness involves strong feelings of concern and worry about the uncertainty surrounding the situation and the impact of the epidemic. Anxiety is the feeling of fear that occurs in threatening or stressful situations, which encourages persons to avoid dangerous places (Dean, 2016). However, if anxiety is overwhelming or persists, it could develop into an anxiety disorder (Dean, 2016). Anxiety disorders seriously affect health and quality of life. A series of symptoms may be experienced, such as worrying all the time, feeling tired, being irritable, struggling to concentrate and sleeping poorly (Dean, 2016). Our results suggested that anxiety was strongly positively correlated with manic depression, fatigue and restlessness (r=0.826, r=0.727, r=0.724, p<0.01). These factors influence cognitive processes and work performance and can even elevate blood pressure and reduce immune system activation (Nechita et al., 2018). The manic depression domain included the original two domains of depression and anger in the AMS. These negative emotions can develop into depressive disorder or even bipolar disorder. Depression can manifest in many ways, such as losing one’s temper, having difficulty falling sleep, being absent-minded, feeling restless, losing one’s appetite or overeating, and even thinking about committing suicide. Depression affects one’s interpersonal relationships, social life, career and sense of self-worth, leading to severe dysfunction (Behere et al., 2017). Bipolar disorder is characterized by several different types of mood episodes, such as depression, mania, hypomania and mixed episodes. There is an increased risk of mortality in individuals with depressive and bipolar disorders (Grande et al., 2016). These disorders should be considered life-threatening.

In this study, sex did not affect the experience of negative emotions except that a greater proportion of women (26.3%) were fatigued. A previous study noted that women were twice as likely to suffer from depression than men and were more likely to have comorbid anxiety (Karger, 2014). Consistent with this observation, Wang et al. (2020) reported higher levels of stress, anxiety, and depression among women in the early stages of the outbreak. However, epidemiological studies further confirmed that COVID-19 predominantly affects men (Huang et al., 2020); therefore, women did not experience as much concern. As a part of the ageing process, the physiological and psychological functions of older adults become weakened. Older adults also experience changes in social roles, social environment, and family circumstances and experience life events such as somatic illness and the death of a spouse. These factors could render older adults more susceptible to geriatric depression (Zhang et al., 2018). In addition, COVID-19 predominantly affects people over 41 years of age, and old age might be associated with increased mortality (Huang et al., 2020); concerns about these risk factors may lead to many serious emotional problems. A higher percentage of elderly people than people under 29 years of age felt tired, restless, anxious, and even depressed. Therefore, we should focus on the group of people over the age of 60 years. This age group had the highest proportion of participants who felt anxious (25%) and had the highest scores for manic depression, restlessness and fatigue (p<0.05). We found that people with no formal education or doctoral degrees had a greater likelihood of experiencing emotional problems and more serious emotional problems during the epidemic. The group with the least education had the largest proportion of people (41.2%) who experienced negative emotions, mainly restlessness (27.5%) and anxiety (21.6%). This might be due to a lack of health information available about COVID-19 (Wang et al., 2020). However, the group with doctoral degrees showed greater degrees of restlessness (p<0.05), manic depression (p>0.05) and anxiety (p>0.05) than the other groups, which was consistent with the literature showing that a high educational level was associated with a greater degree of fear during the SARS outbreak (Wu et al., 2009). In the analysis stratified by occupation, our findings showed that during the outbreak, people in all occupations were affected by negative emotion. However, healthcare workers were not more likely to feel negative emotions and did not feel negative emotions more strongly than the other occupational groups, even though they were at the greatest risk of infection among people with different occupations. This result was different from that of Lai et al. (2020) at the end of January, which showed that more than half of healthcare workers were depressed and distressed. We believe that altruism may help protect some healthcare workers against these negative impacts (Wu et al., 2009). Meanwhile, the rapid deployment of a comprehensive prevention program was initiated by the government as soon as possible, which could have decreased anxiety and depression levels (Chen et al., 2006). In addition, we found that 6% of students felt higher levels of manic depression than teachers (p<0.01), but this percentage was a relatively low compared with the 24.9% of college students who reported anxiety in a study by Cao et al. (2020). These findings suggest that the development of online teaching to control the epidemic decreased the negative impact on students who worried about a delay in their studies and future employment. Employees experienced anxiety more strongly than teachers (p<0.05), which might have been because they were more worried about the financial losses caused by the epidemic (Yuan et al., 2020). It was very surprising that although Hubei was the main site of the outbreak, Hubei did not have the largest proportion of people reporting negative emotions, which may be due to the effective and rapid government control. However, people in Hubei Province still had the highest proportion of people who experienced serious negative emotions (manic depression and anxiety), and people from Hubei Province also had the strongest degree of these emotions; these findings were significantly correlated with the number of confirmed cases or deaths (p<0.05).

Our study also has some limitations that should be considered. The survey was carried out in the late stage of the epidemic when the epidemic had been effectively contained and people’s negative emotions may have been substantially relieved. This timing may have affected the results of the survey. The conclusions would be enhanced by the inclusion of more participants and a longer study duration.

## Conclusions

We present the following conclusions and recommendations based on this study. First, emotional problems occurred in 33.9% of uninfected people. People who reported emotional problems experienced significantly more fatigue, restlessness, anxiety and depression than those who did not report emotional problems; decreased happiness; and a lack of vigour. Second, we should pay attention to special groups of people, such as people over 60 years old, those with doctoral degrees and those employed by private companies, as these people are more likely to feel serious negative emotions, such as anxiety and depression. Third, the perception of negative emotions was not more prominent among healthcare workers than among other occupational groups in terms of the proportion of people who reported negative emotions or the degree to which they were experienced; however, approximately 30% of healthcare workers were affected by the epidemic. We should pay attention to the emotional states of healthcare workers because of their high risk of infection. Fourth, the uninfected people in the areas most affected by the pandemic reported relatively serious emotional problems, and psychological care or treatment should be considered. Finally, the effective control of the number of confirmed cases and deaths will significantly reduce the public perception of negative emotions. Meanwhile, psychological guidance and interventions should be provided as early as possible to prevent subsequent serious consequences for people with initially mild emotional problems since negative emotions are significantly correlated with each other. Moreover, the modified scale had good reliability and validity and could be used to survey the mood states of the public during public health events to enable officials to detect problems and intervene early.

## Data Availability

All data produced in the present study are available upon reasonable request to the authors.

## Acknowledgements

The authors express sincere gratitude to Rui Chen and Jason Dong for modifying the description of the AMS in Chinese.

---

**Supplementary Table 1.** Characteristics of the survey participants

|  |  | Number | Percentage (%) |
| --- | --- | --- | --- |
| **Gender** | Male | 301 | 53.1 |
|  | Female | 266 | 46.9 |
| **Age range, years** | ≤29 | 98 | 17.3 |
|  | 30-39 | 267 | 47.1 |
|  | 40-49 | 113 | 19.9 |
|  | 50-59 | 53 | 9.4 |
|  | ≥60 | 36 | 6.3 |
| **Education level** | Junior college | 66 | 11.6 |
|  | Bachelor's | 226 | 39.9 |
|  | Master's | 126 | 22.2 |
|  | Doctoral | 98 | 17.3 |
|  | Other | 51 | 9 |
| **Occupation** | Medical staff | 136 | 24 |
|  | Teacher | 85 | 15 |
|  | Student | 50 | 8.8 |
|  | Public official | 29 | 5.1 |
|  | Employee | 115 | 20.3 |
|  | Other | 152 | 26.8 |
| **Location** | Hubei | 79 | 13.9 |
|  | Guangdong | 17 | 3 |
|  | Chongqing | 126 | 22.2 |
|  | Sichuan | 136 | 24 |
|  | Shanghai | 26 | 4.6 |
|  | Jiangsu | 31 | 5.5 |
|  | Guizhou | 14 | 2.5 |
|  | Beijing | 31 | 5.5 |
|  | Other | 107 | 18.8 |

(Other places included Henan, 9; Hunan, 6; Zhejiang, 10; Yunnan, 1; Xinjiang, 3;
Xizang, 1; Tianjin, 9; Shanxi, 6; Shandong, 13; Qinghai, 1; Ningxia, 1; Neimenggu, 2;
Liaoning, 12; Jiangxi, 3; Jilin 1; Heilongjiang, 1; Hebei, 8; Hainan, 2; Guangxi, 2;
Gansu, 2; Fujian, 7; and Anhui, 7)

**Supplementary Table 2.** Correlation matrix of the six emotions

|  | **Manic depression** | **Happiness** | **Restlessness** | **Vigour** | **Fatigue** | **Anxiety** |
| --- | --- | --- | --- | --- | --- | --- |
| Manic depression | 1 |  |  |  |  |  |
| Happiness | -0.307** | 1 |  |  |  |  |
| Restlessness | 0.742** | -0.327** | 1 |  |  |  |
| Vigour | -0.376** | 0.543** | -0.332** | 1 |  |  |
| Fatigue | 0.737** | -0.348** | 0.596** | -0.504** | 1 |  |
| Anxiety | 0.826** | -0.339** | 0.724** | -0.353** | 0.727** | 1 |

\*\*P<0.01

**Supplementary Table 3.** Characteristics of respondents with negative emotions (n=192)

| Variable | Number | Percentage |
| --- | --- | --- |
| **Age, years** |  |  |
| Mean (SD) | 38.69(11.03) |  |
| Range | 20-71 |  |
| ≤29 | 31 | 31.6% |
| 30-39 | 96 | 36.0% |
| 40-49 | 36 | 31.9% |
| 50-59 | 17 | 32.0% |
| ≥60 | 12 | 33.3% |
| **Gender** |  |  |
| Male | 101 | 33.6% |
| Female | 91 | 34.2% |
| **Education level** |  |  |
| Junior college | 18 | 27.3% |
| Bachelor's | 78 | 34.5% |
| Master's | 39 | 31.0% |
| Doctoral | 36 | 36.7% |
| Other | 21 | 41.2% |
| **Occupation** |  |  |
| Medical staff | 41 | 30.1% |
| Teacher | 32 | 37.6% |
| Student | 17 | 34.0% |
| Public official | 10 | 34.5% |
| Employee | 40 | 34.8% |
| Other | 52 | 34.2% |
| **Location (areas with samples smaller than 15 were not analysed)** |  |  |
| Hubei | 29 | 36.7% |
| Sichuan | 43 | 31.6% |
| Chongqing | 40 | 31.7% |
| Beijing | 14 | 45.2% |
| Jiangsu | 8 | 25.8% |
| Shanghai | 11 | 42.3% |
| Guangdong | 5 | 29.4% |
| Guizhou | 4 | 28.6% |

The percentage (%) = the number of people in the negative emotion group/the number of people in this group in the total sample.

**Supplementary Table 4.**
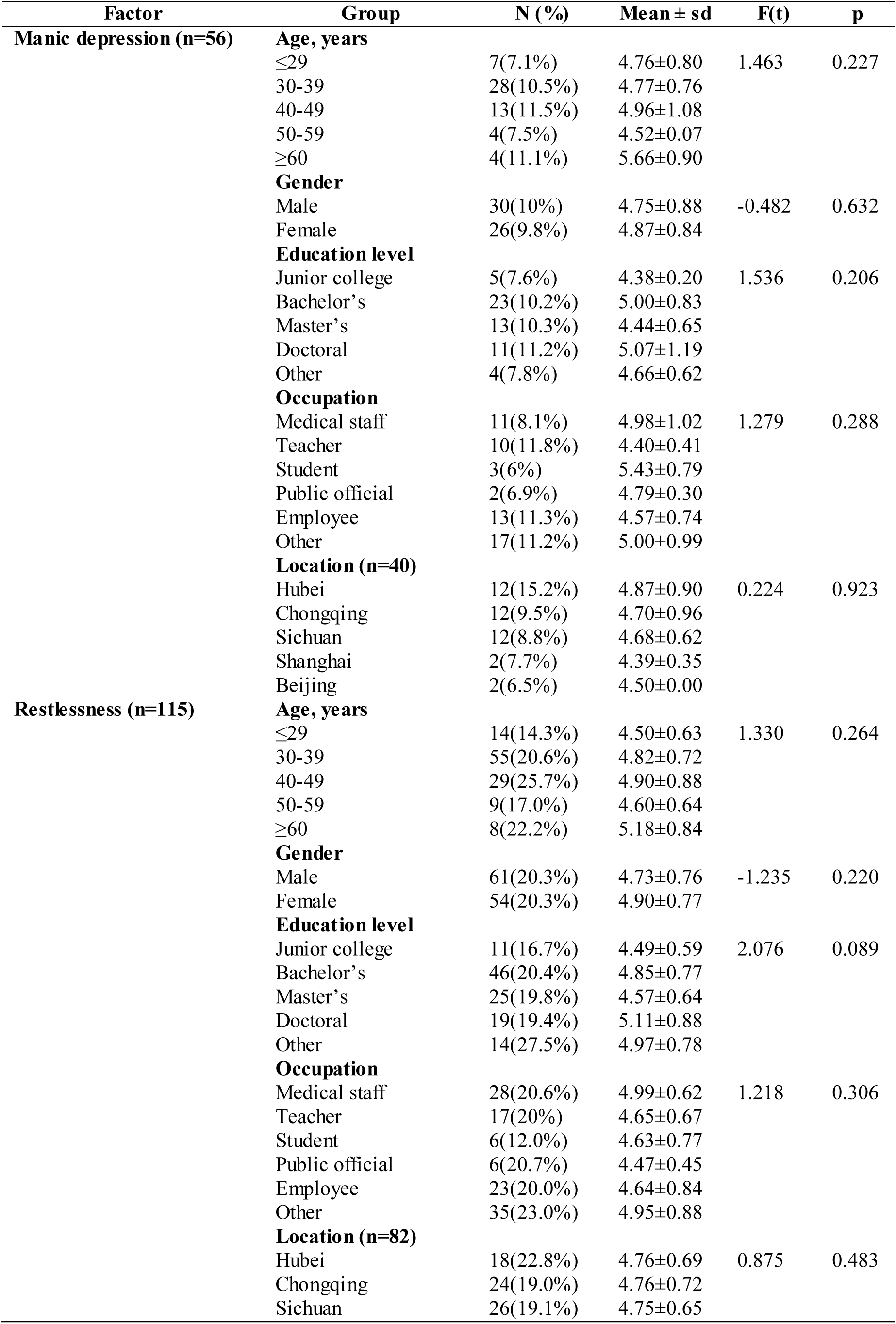
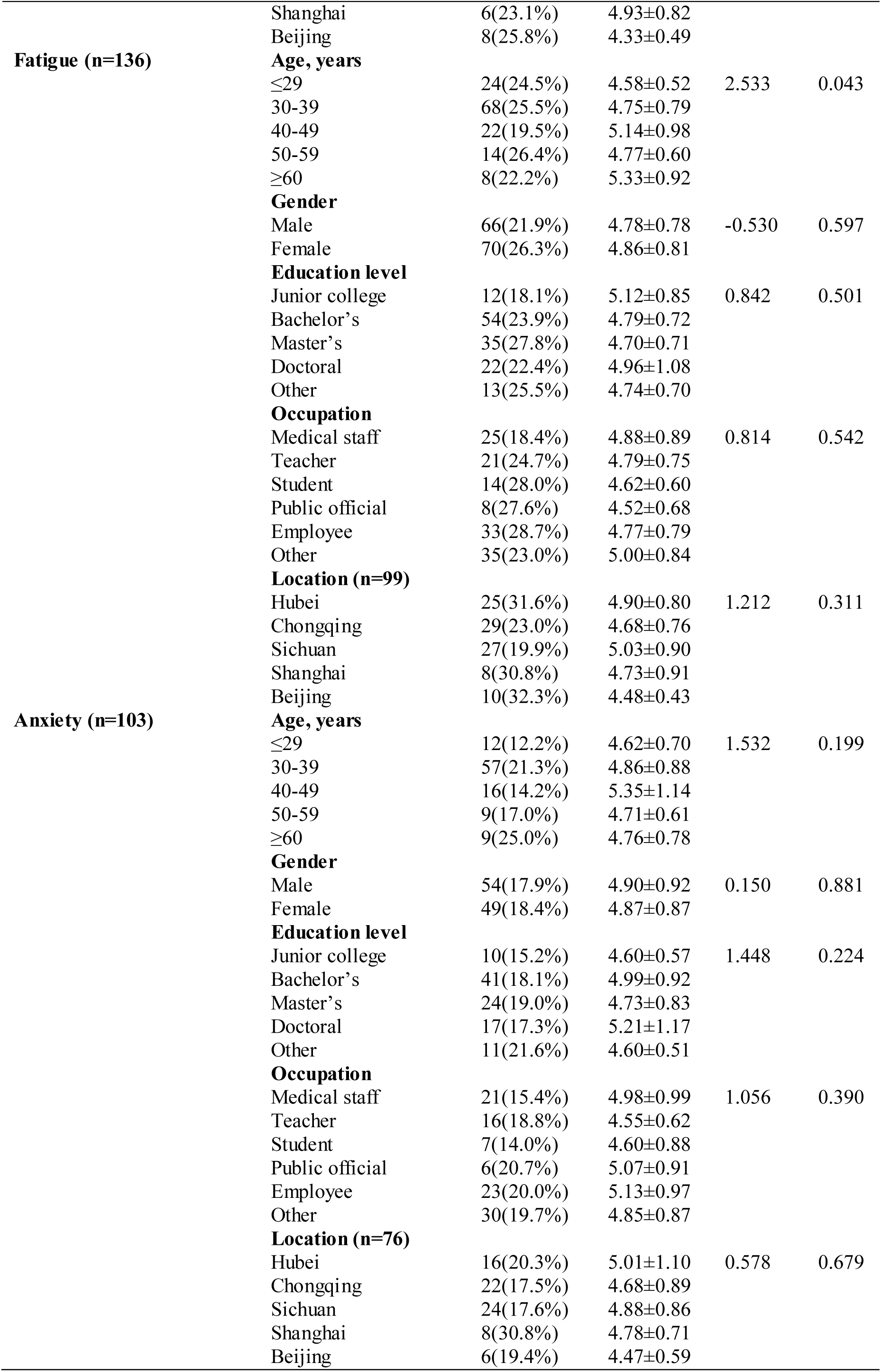
Characteristics of the different groups and comparison of negative mood perceptions among them

**Supplementary Table 5.** Correlation matrix of cumulative cases, total deaths and the six emotions (n=137)

|  | **Cumulative cases** | **Total deaths** | **Manic depression** | **Happiness** | **Restlessness** | **Vigour** | **Fatigue** | **Anxiety** |
| --- | --- | --- | --- | --- | --- | --- | --- | --- |
| **Cumulative cases** | 1 | 1.000** | 0.230** | -0.128 | 0.075 | -0.234** | 0.219** | 0.170* |
| **Total deaths** | 1.000** | 1 | 0.230** | -0.127 | 0.074 | -0.234** | 0.219** | 0.169* |

The number of confirmed cases and deaths from the provincial health commissions
(Hubei, Chongqing, Sichuan, Shanghai and Beijing) was updated at 24:00 on March 16,
2020. \*\* P<0.01, \* P<0.05.

